# The Association of Cardiovascular Comorbidities with Malignant and Benign Colorectal Neoplasms

**DOI:** 10.64898/2026.08.04.26359655

**Authors:** Farah Wani, Ivan Marrufo, Vedang Bhavsar, Rebecca Whitmer, Jagmeet Singh, Asim Kichloo

**Affiliations:** Assistant Professor, Department of Family and Osteopathic Medicine, Texas College of Osteopathic Medicine, University of North Texas Health Sciences Center, Fort Worth, TX 76107; Medical Student, Texas College of Osteopathic Medicine, University of North Texas Health Sciences Center, Fort Worth, TX 76107; Medical Director of Nuclear and Echovascular Imaging, Northeast Georgia Medical Center, Assistant Professor of Medicine, Augusta University, Gainesville, GA 30501; Resident Physician, Texas Health Harris Methodist Fort Worth, TX 76104; Associate Professor, Department of Internal Medicine, Guthrie Robert Packet Hospital, Sayre, PA 18840; Chair Professor, Department of Internal Medicine, Texas College of Osteopathic Medicine, University of North Texas Health Sciences Center, 3500 Camp Bowie Blvd, Fort Worth, TX 76107

**Keywords:** Lower GI bleed, Antiplatelet Therapy, Oral Anticoagulation Therapy, Colorectal neoplasms

## Abstract

**Background:** Each year, thousands of patients are diagnosed with gastrointestinal (GI) bleeding. Many of these patients undergo colonoscopy and are subsequently diagnosed with benign or malignant colorectal neoplasms. The aim of this study is to evaluate whether patients with cardiovascular comorbidities, many of whom are on anti-platelet or anticoagulation therapy, are more likely to be diagnosed with new benign or malignant colorectal neoplasms than patients without cardiovascular comorbidities.

**Methods:** From the 2007–2011 NIS, 802,080 primary lower GI bleed admissions (ICD-9) were identified; 283,925 had cardiovascular comorbidities. We compared patients with vs without these comorbidities for the primary outcome of new benign or malignant colorectal neoplasms. Weighted analyses accounting for the complex survey design were performed in SAS 9.4, with trends assessed using Cochran–Armitage tests and linear regression and associations with colorectal cancer stage evaluated by multinomial logistic regression (p ≤ 0.05).

**Results:** Between 2007-2011, the odds of malignant colorectal neoplasms increased between 49.9% and 65.3% for patients without cardiovascular comorbidities when presenting with a lower GI bleed, compared to those with cardiovascular comorbidities. Between 2007 and 2011, the most notable and statistically significant difference was observed in 2010, when patients with cardiovascular comorbidities had 10.5% higher odds of being diagnosed with benign colorectal neoplasms than those without cardiovascular comorbidities.

**Conclusion:** Patients presenting with a lower gastrointestinal bleed without cardiovascular comorbidities were significantly more likely to be diagnosed with malignant colorectal neoplasms than those with cardiovascular comorbidities.

## Introduction

In the United States, colorectal cancer (CRC) is the third most prevalent cancer, after only breast and lung cancer.^1^ CRC is also the second highest cause of mortality, after only lung cancer.^1^ Although most patients with colorectal carcinoma may be asymptomatic or have nonspecific symptoms, such as fatigue from iron deficiency anemia or abdominal pain, the most common presenting symptom for colorectal carcinoma that leads to medical attention is lower gastrointestinal (GI) bleeding.^2,3^ A lower GI bleed is defined as any bleeding distal to the ileocecal valve and most commonly presents as hematochezia.^4^ Less commonly, if blood has been in the GI tract for a significant amount of time, a lower GI bleed may present as melena.^5^ Due to the high incidence and mortality associated with Colorectal Cancer (CRC), patients with lower GI bleeding require prompt screening to rule out CRC.

Lower GI bleeding represents nearly 20% of all GI bleeding.^6^ It has a broad differential, including diverticulosis, mesenteric ischemia, inflammatory bowel disease, and arteriovenous malformations.^7^ In comparison, CRC is a significantly less common cause of lower GI bleeding. In fact, the incidence of CRC in the United States is 38.7 per 100,000 people - a rate that has been declining by approximately 2% annually.^8,9^ However, despite decreasing incidences, the 5-year survival rate for CRC overall remains only 65%, dropping sharply to a bleak 12% in late-stage CRC (Stage IV).^10^

There are both modifiable and non-modifiable risk factors for CRC, with notable disparities across different populations. The risk of developing CRC is approximately twice as high in men than in women and increases with age.^11–13^ Specific hereditary conditions confer a higher risk, such as Familial Adenomatous Polyposis (FAP), Lynch Syndrome, and those with a family history of CRC.^14^ Patients who have history of inflammatory bowel disease, such as Ulcerative Colitis and Crohn’s Disease, are also at increased risk of developing CRC compared to the general population.^15^ With regards to modifiable risk factors, CRC incidence is higher among individuals with high fat intake, consumption of red or processed meats, smoking history, and alcohol use.^16^

Screening for CRC has been shown to reduce mortality by approximately 50% with current screening rates.^17^ In fact, a significant portion CRC mortality has been attributed directly to lack of screening.^18^ There are various methods for CRC screening including stool-based tests and direct visualization methods; however, colonoscopy with biopsy remains the gold standard due to its superior ability to reduce mortality.^19^ It is currently recommended to start colon cancer screening in the general population at the age of 45 for both men and women, with earlier ages adjusted accordingly for high risk individuals, such as those with family history of CRC, Lynch Syndrome, and FAP.^17^

Although it is known that antiplatelet therapy and oral anticoagulation therapy increase the risk of bleeding, there is a surprising lack of data about whether the cardiovascular comorbidities that are often treated with these medications affect the detection of CRC.^20,21^ This cross-sectional study is the first to analyze National Inpatient Sample (NIS) data to examine whether patients with cardiovascular comorbidities have different odds of being diagnosed with benign colorectal neoplasms and malignant CRC. We hypothesize that patients with cardiovascular comorbidities may have higher odds of benign colorectal neoplasms and lower odds of malignant colorectal cancer; however, because the NIS database does not include outpatient medication data, early bleeding due to antiplatelet or anticoagulant therapy, is only a proposed mechanism, which will need further medication-specific studies to fully evaluate.

## Methods

### Data Source

The NIS is the largest publicly available all-payer inpatient health care database in the United States. It is part of the Healthcare Cost and Utilization Project (HCUP) and is maintained by the Agency for Health Care Quality and Research (AHQR). It is one of the most useful databases for studying outcomes and trends of various procedures and diseases. It is comprised of de-identified data collected from 20% of community hospitals in 46 states across the United States. Each hospitalization is representative of one primary diagnosis, up to 29 secondary diagnoses, and up to 15 procedures, all coded using the International Clinical Modification Codes (ICD9 and ICD10). The data includes information regarding admission status, patient demographics, admitting diagnosis, comorbidities, healthcare facility status (rural vs. urban), discharge diagnosis, outcomes, length of stay, and cost during hospitalization.

Patients were filtered using International Classification of Diseases, ninth revision, and clinical modification codes (ICD-9 Codes). In this cross-sectional study using NIS data from 2007 and 2011, we studied all hospitalizations with a diagnosis of lower GI bleeding. The GI bleeding was identified with validated International Classification of Diseases, 9th Revision, Clinical Modification code 578.1. Patient characteristics considered for those with lower GI bleeding included age, sex, race, region, location, median household income, payment, source of admission, type of admission, disposition status, the history of obesity, hypertension, diabetes mellitus, congestive heart failure, chronic pulmonary disease, renal failure, neurological disorders, anemia, coagulopathy disorders, and depression. Malignant colorectal neoplasms were identified with the validated ICD-9 codes 153.0, 153.1, 153.2, 153.3, 153.4, 153.5, 153.6, 153.7, 153.8, 153.9, 154.0, 154.1, 154.2, 154.3, and 154.8. Benign colorectal neoplasms were identified with the validated ICD-9 codes 211.3 and 211.4. The cardiovascular comorbidities that most commonly require antiplatelet therapy, anticoagulation, or both were also considered. Atrial fibrillation and flutter were identified with the validated ICD-9 codes 427.31 and 427.32; ischemic stroke with the validated ICD-9 codes 434.00, 434.01, 434.10, 434.11, 434.90, and 434.91; peripheral arterial disease (PAD) with the validated ICD-9 codes 440.20, 440.21, 440.22, 440.23, 440.24, and 440.29; coronary arterial disease (CAD) with the validated ICD-9 codes 414.0, 414.01, 414.02, 414.03, 414.04, 414.05, 414.06, and 414.07; and valvular heart disease (arteriosclerosis of valves) with the validated ICD-9 codes 424.99, 424.1, 424.0, 424.3, and 424.2. Notably, deep vein thrombosis and pulmonary embolus were excluded as comorbidities. Although these conditions may require antiplatelet or anticoagulation therapy, the duration of therapy is often shorter and more variable than the other comorbidities considered. Additionally, it is impossible to distinguish between provoked and unprovoked clots within the NIS database, which significantly affects clinical management.

### Primary Outcomes

Our objectives were to analyze the following:

- To determine the odds of newly diagnosed malignant and benign colorectal neoplasms in an inpatient setting, for patients with the defined cardiovascular comorbidities presenting with lower GI bleeding, versus the odds of newly diagnosed malignant and benign colorectal neoplasms in those patients without cardiovascular comorbidities.
- To determine the odds of newly diagnosed *malignant only* colorectal neoplasms in an inpatient setting, for patients with cardiovascular comorbidities presenting with lower GI bleeding, versus the odds of newly diagnosed *malignant only* colorectal neoplasms in those patients without cardiovascular comorbidities.
- To determine the odds of newly diagnosed *benign only* colorectal neoplasms in an inpatient setting, for patients with cardiovascular comorbidities presenting with lower GI bleeding, versus the odds of newly diagnosed *benign only* colorectal neoplasms in those patients without cardiovascular comorbidities.
- To determine the odds of malignant colorectal neoplasms versus the odds of benign colorectal neoplasms diagnosed in patients with cardiovascular comorbidities.

### Statistical Analysis

To account for weights in the stratified survey design, the SAS 9.4 (SAS Institute Inc, Cary, NC) was used for the statistical analyses. The weights were applied in the statistical estimation process by incorporating the variables for strata (used to post-stratify hospital), clusters, (HCUP hospital identification number), and discharge weight, (to representing how many discharges it represents in the full U.S. hospital population). Descriptive statistics included mean with standard deviation and quantiles for continuous variables and counts with percentages for categorical variables. To test for trends in proportions, the Cochrane Armitage test was implemented. The trend test for continuous variables was obtained by using simple linear regression. The (multinomial) logistics regression was adopted to examine the association between variables. Logistic regression analyses were performed using age, sex, cardiovascular comorbidity status as independent variables. Additional demographics and socioeconomic characteristics available in the NIS database, including race/ethnicity, geographic region, income quartile, and insurance status, were not incorporated in the regression models. All the analytical results were considered to be significant when p-values were less than or equal to 0.05.

## Results

We identified a total of 151,123 hospitalizations for lower GI bleeding for the year 2007, and 177,930, 159,968, 161,649, and 151,410 hospitalizations for the years 2008, 2009, 2010, and 2011, respectively. Of these hospitalizations, an average of 35.40% of patients had at least one of the pertinent cardiovascular comorbidities, including atrial fibrillation (AF), cardiovascular accident (CVA), peripheral arterial disease (PAD), coronary artery disease (CAD), valvular heart disease (VHD), or any combination of these comorbidities (Table 1). Table 1 further demonstrates the demographic data of the study population.

**Table 1:**
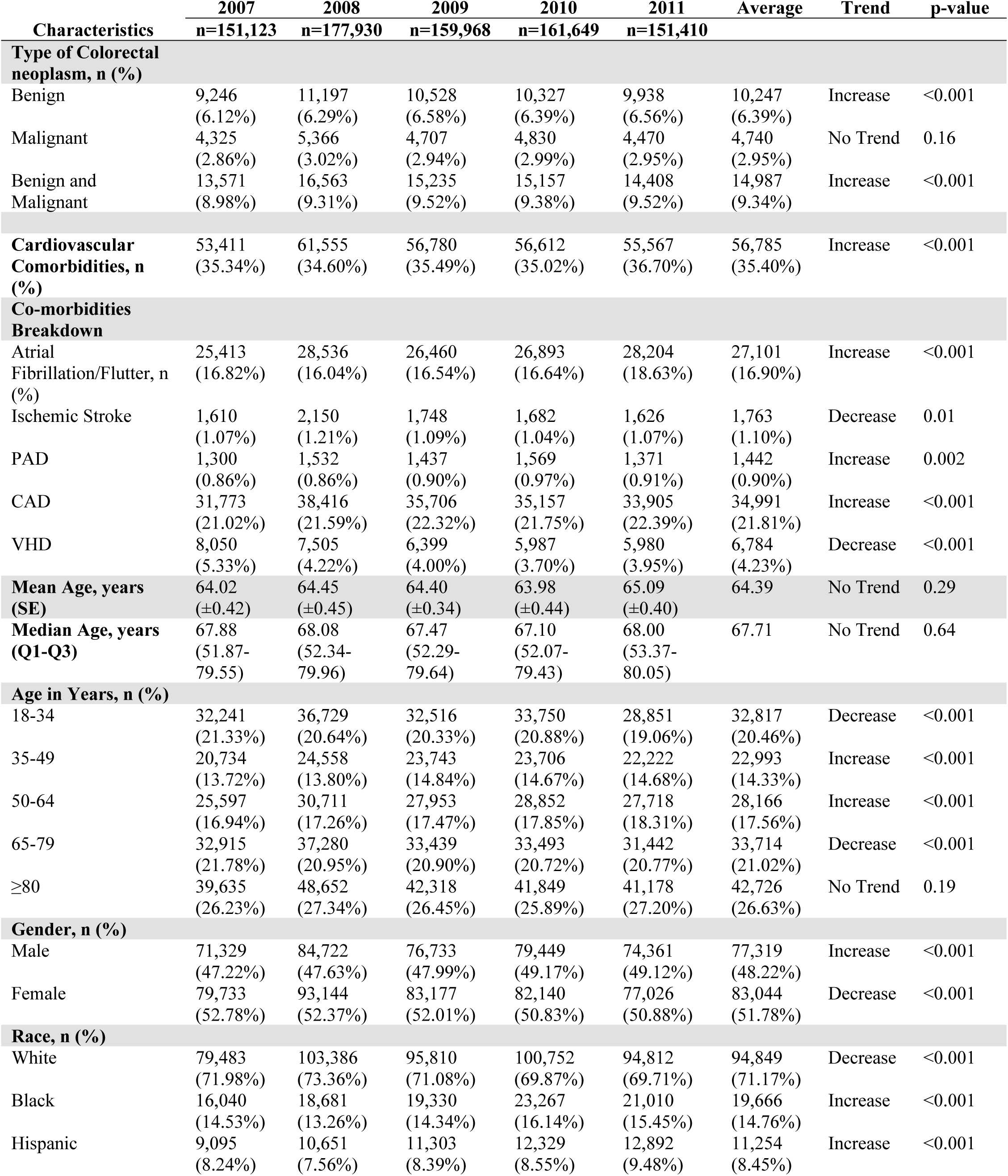
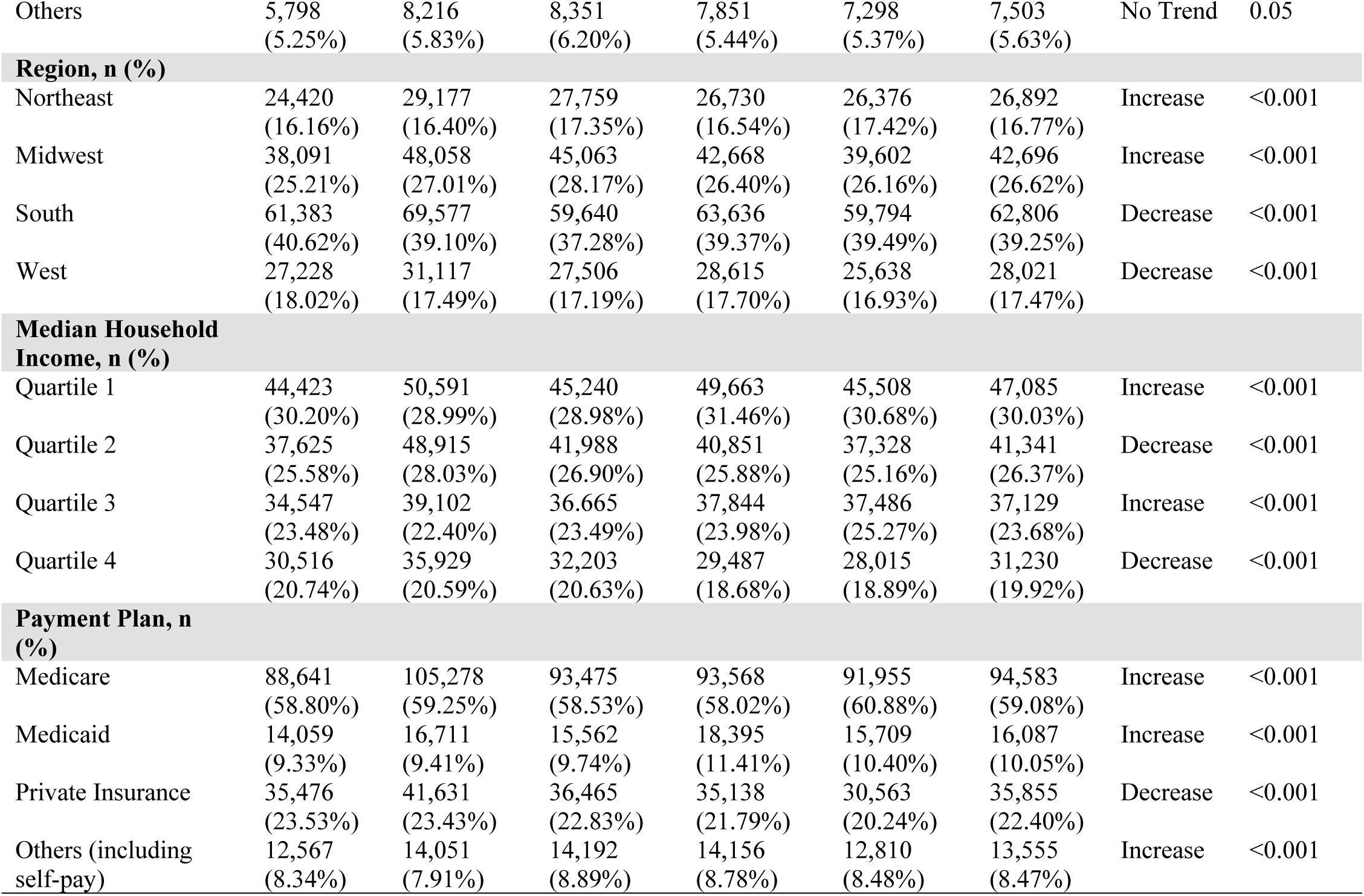
Demographics. Demographic and baseline characteristics of the study population from the National Inpatient Sample (NIS) database.

| Characteristics | 2007<br>n=151,123 | 2008<br>n=177,930 | 2009<br>n=159,968 | 2010<br>n=161,649 | 2011<br>n=151,410 | Average | Trend | p-value |
| --- | --- | --- | --- | --- | --- | --- | --- | --- |
| **Type of Colorectal neoplasm, n (%)** |  |  |  |  |  |  |  |  |
| Benign | 9,246<br>(6.12%) | 11,197<br>(6.29%) | 10,528<br>(6.58%) | 10,327<br>(6.39%) | 9,938<br>(6.56%) | 10,247<br>(6.39%) | Increase | <0.001 |
| Malignant | 4,325<br>(2.86%) | 5,366<br>(3.02%) | 4,707<br>(2.94%) | 4,830<br>(2.99%) | 4,470<br>(2.95%) | 4,740<br>(2.95%) | No Trend | 0.16 |
| Benign and Malignant | 13,571<br>(8.98%) | 16,563<br>(9.31%) | 15,235<br>(9.52%) | 15,157<br>(9.38%) | 14,408<br>(9.52%) | 14,987<br>(9.34%) | Increase | <0.001 |
| **Cardiovascular Comorbidities, n (%)** |  |  |  |  |  |  |  |  |
| Cardiovascular Comorbidities, n (%) | 53,411<br>(35.34%) | 61,555<br>(34.60%) | 56,780<br>(35.49%) | 56,612<br>(35.02%) | 55,567<br>(36.70%) | 56,785<br>(35.40%) | Increase | <0.001 |
| **Co-morbidities Breakdown** |  |  |  |  |  |  |  |  |
| Atrial Fibrillation/Flutter, n (%) | 25,413<br>(16.82%) | 28,536<br>(16.04%) | 26,460<br>(16.54%) | 26,893<br>(16.64%) | 28,204<br>(18.63%) | 27,101<br>(16.90%) | Increase | <0.001 |
| Ischemic Stroke | 1,610<br>(1.07%) | 2,150<br>(1.21%) | 1,748<br>(1.09%) | 1,682<br>(1.04%) | 1,626<br>(1.07%) | 1,763<br>(1.10%) | Decrease | 0.01 |
| PAD | 1,300<br>(0.86%) | 1,532<br>(0.86%) | 1,437<br>(0.90%) | 1,569<br>(0.97%) | 1,371<br>(0.91%) | 1,442<br>(0.90%) | Increase | 0.002 |
| CAD | 31,773<br>(21.02%) | 38,416<br>(21.59%) | 35,706<br>(22.32%) | 35,157<br>(21.75%) | 33,905<br>(22.39%) | 34,991<br>(21.81%) | Increase | <0.001 |
| VHD | 8,050<br>(5.33%) | 7,505<br>(4.22%) | 6,399<br>(4.00%) | 5,987<br>(3.70%) | 5,980<br>(3.95%) | 6,784<br>(4.23%) | Decrease | <0.001 |
| Mean Age, years (SE) | 64.02<br>(±0.42) | 64.45<br>(±0.45) | 64.40<br>(±0.34) | 63.98<br>(±0.44) | 65.09<br>(±0.40) | 64.39 | No Trend | 0.29 |
| Median Age, years (Q1-Q3) | 67.88<br>(51.87-79.55) | 68.08<br>(52.34-79.96) | 67.47<br>(52.29-79.64) | 67.10<br>(52.07-79.43) | 68.00<br>(53.37-80.05) | 67.71 | No Trend | 0.64 |
| **Age in Years, n (%)** |  |  |  |  |  |  |  |  |
| 18-34 | 32,241<br>(21.33%) | 36,729<br>(20.64%) | 32,516<br>(20.33%) | 33,750<br>(20.88%) | 28,851<br>(19.06%) | 32,817<br>(20.46%) | Decrease | <0.001 |
| 35-49 | 20,734<br>(13.72%) | 24,558<br>(13.80%) | 23,743<br>(14.84%) | 23,706<br>(14.67%) | 22,222<br>(14.68%) | 22,993<br>(14.33%) | Increase | <0.001 |
| 50-64 | 25,597<br>(16.94%) | 30,711<br>(17.26%) | 27,953<br>(17.47%) | 28,852<br>(17.85%) | 27,718<br>(18.31%) | 28,166<br>(17.56%) | Increase | <0.001 |
| 65-79 | 32,915<br>(21.78%) | 37,280<br>(20.95%) | 33,439<br>(20.90%) | 33,493<br>(20.72%) | 31,442<br>(20.77%) | 33,714<br>(21.02%) | Decrease | <0.001 |
| ≥80 | 39,635<br>(26.23%) | 48,652<br>(27.34%) | 42,318<br>(26.45%) | 41,849<br>(25.89%) | 41,178<br>(27.20%) | 42,726<br>(26.63%) | No Trend | 0.19 |
| **Gender, n (%)** |  |  |  |  |  |  |  |  |
| Male | 71,329<br>(47.22%) | 84,722<br>(47.63%) | 76,733<br>(47.99%) | 79,449<br>(49.17%) | 74,361<br>(49.12%) | 77,319<br>(48.22%) | Increase | <0.001 |
| Female | 79,733<br>(52.78%) | 93,144<br>(52.37%) | 83,177<br>(52.01%) | 82,140<br>(50.83%) | 77,026<br>(50.88%) | 83,044<br>(51.78%) | Decrease | <0.001 |
| **Race, n (%)** |  |  |  |  |  |  |  |  |
| White | 79,483<br>(71.98%) | 103,386<br>(73.36%) | 95,810<br>(71.08%) | 100,752<br>(69.87%) | 94,812<br>(69.71%) | 94,849<br>(71.17%) | Decrease | <0.001 |
| Black | 16,040<br>(14.53%) | 18,681<br>(13.26%) | 19,330<br>(14.34%) | 23,267<br>(16.14%) | 21,010<br>(15.45%) | 19,666<br>(14.76%) | Increase | <0.001 |
| Hispanic | 9,095<br>(8.24%) | 10,651<br>(7.56%) | 11,303<br>(8.39%) | 12,329<br>(8.55%) | 12,892<br>(9.48%) | 11,254<br>(8.45%) | Increase | <0.001 |
| Others | 5,798<br>(5.25%) | 8,216<br>(5.83%) | 8,351<br>(6.20%) | 7,851<br>(5.44%) | 7,298<br>(5.37%) | 7,503<br>(5.63%) | No Trend | 0.05 |
| **Region, n (%)** |  |  |  |  |  |  |  |  |
| Northeast | 24,420<br>(16.16%) | 29,177<br>(16.40%) | 27,759<br>(17.35%) | 26,730<br>(16.54%) | 26,376<br>(17.42%) | 26,892<br>(16.77%) | Increase | <0.001 |
| Midwest | 38,091<br>(25.21%) | 48,058<br>(27.01%) | 45,063<br>(28.17%) | 42,668<br>(26.40%) | 39,602<br>(26.16%) | 42,696<br>(26.62%) | Increase | <0.001 |
| South | 61,383<br>(40.62%) | 69,577<br>(39.10%) | 59,640<br>(37.28%) | 63,636<br>(39.37%) | 59,794<br>(39.49%) | 62,806<br>(39.25%) | Decrease | <0.001 |
| West | 27,228<br>(18.02%) | 31,117<br>(17.49%) | 27,506<br>(17.19%) | 28,615<br>(17.70%) | 25,638<br>(16.93%) | 28,021<br>(17.47%) | Decrease | <0.001 |
| **Median Household Income, n (%)** |  |  |  |  |  |  |  |  |
| Quartile 1 | 44,423<br>(30.20%) | 50,591<br>(28.99%) | 45,240<br>(28.98%) | 49,663<br>(31.46%) | 45,508<br>(30.68%) | 47,085<br>(30.03%) | Increase | <0.001 |
| Quartile 2 | 37,625<br>(25.58%) | 48,915<br>(28.03%) | 41,988<br>(26.90%) | 40,851<br>(25.88%) | 37,328<br>(25.16%) | 41,341<br>(26.37%) | Decrease | <0.001 |
| Quartile 3 | 34,547<br>(23.48%) | 39,102<br>(22.40%) | 36,665<br>(23.49%) | 37,844<br>(23.98%) | 37,486<br>(25.27%) | 37,129<br>(23.68%) | Increase | <0.001 |
| Quartile 4 | 30,516<br>(20.74%) | 35,929<br>(20.59%) | 32,203<br>(20.63%) | 29,487<br>(18.68%) | 28,015<br>(18.89%) | 31,230<br>(19.92%) | Decrease | <0.001 |
| **Payment Plan, n (%)** |  |  |  |  |  |  |  |  |
| Medicare | 88,641<br>(58.80%) | 105,278<br>(59.25%) | 93,475<br>(58.53%) | 93,568<br>(58.02%) | 91,955<br>(60.88%) | 94,583<br>(59.08%) | Increase | <0.001 |
| Medicaid | 14,059<br>(9.33%) | 16,711<br>(9.41%) | 15,562<br>(9.74%) | 18,395<br>(11.41%) | 15,709<br>(10.40%) | 16,087<br>(10.05%) | Increase | <0.001 |
| Private Insurance | 35,476<br>(23.53%) | 41,631<br>(23.43%) | 36,465<br>(22.83%) | 35,138<br>(21.79%) | 30,563<br>(20.24%) | 35,855<br>(22.40%) | Decrease | <0.001 |
| Others (including self-pay) | 12,567<br>(8.34%) | 14,051<br>(7.91%) | 14,192<br>(8.89%) | 14,156<br>(8.78%) | 12,810<br>(8.48%) | 13,555<br>(8.47%) | Increase | <0.001 |

Using this data, we aimed to achieve statistical validation of trends related to CRC that are widely recognized in the literature. For example, we first confirmed that the odds of CRC increase with age among patients presenting with lower GI bleeding (Table 2). Based on NIS data from 2007 to 2011, among patients presenting with lower GI bleeding, the odds of having malignant CRC or benign colorectal neoplasms increased by 1.2% to 1.7% with each additional year of age. We also confirmed males have a higher chance of developing CRC compared to females (Table 2). Among patients presenting with lower GI bleeding from 2007 to 2011, males had 22.9% to 37.8% higher odds of developing either malignant or benign colorectal neoplasms than females. After validating these findings, we further confirmed that malignant CRC and benign colorectal neoplasms demonstrated similar increases in odds within the same patient population (Tables 3 and 4).

**Table 2:** All Colorectal Neoplasms. The odds ratios (OR) percentage of having colorectal neoplasms (including malignant OR benign) (reference=no neoplasms), using age and gender (reference=female) as independent variables. There is sufficient evidence to conclude the odds of having malignant or benign colorectal neoplasms increases by 1.2 - 1.7% for each additional year of age. Furthermore, males have 22.9-37.8% higher odds of having malignant or benign colorectal neoplasms than females. The highlighted values are those that are statistically significant.

|  | 2007<br>Odds Ratio<br>Percentage<br>(p-value) | 2008<br>Odds Ratio<br>Percentage<br>(p-value) | 2009<br>Odds Ratio<br>Percentage<br>(p-value) | 2010<br>Odds Ratio<br>Percentage<br>(p-value) | 2011<br>Odds Ratio<br>Percentage<br>(p-value) |
| --- | --- | --- | --- | --- | --- |
| **Age** | 1.7% (<0.001) | 1.6% (<0.001) | 1.5% (<0.001) | 1.4% (<0.001) | 1.2% (<0.001) |
| **Gender** | 37.8% (<0.001) | 22.9% (<0.001) | 37.5% (<0.001) | 24.6% (<0.001) | 32.7% (<0.001) |

**Table 3:** Malignant Colorectal neoplasms. The odds ratios (OR) percentage of having malignant neoplasms only (reference= benign or no neoplasms), using age and gender (reference=female) as independent variables. Among patients with lower gastrointestinal (GI) bleeding, there is sufficient evidence to conclude the odds of having malignant colorectal neoplasms increases by 1.8% - 2.3% for each additional year of age. Furthermore, males have 17.1-46.9% higher odds than females of having malignant colorectal neoplasms. The highlighted values are those that are statistically significant.

|  | 2007<br>Odds Ratio<br>Percentage<br>(p-value) | 2008<br>Odds Ratio<br>Percentage<br>(p-value) | 2009<br>Odds Ratio<br>Percentage<br>(p-value) | 2010<br>Odds Ratio<br>Percentage<br>(p-value) | 2011<br>Odds Ratio<br>Percentage<br>(p-value) |
| --- | --- | --- | --- | --- | --- |
| Age | 2.3% (<0.001) | 2.2% (<0.001) | 2.2% (<0.001) | 2.2% (<0.001) | 1.8% (<0.001) |
| Gender | 46.9% (<.001) | 17.1% (0.03) | 42.5% (<.001) | 19.0% (0.009) | 38.6% (<.001) |

**Table 4:**
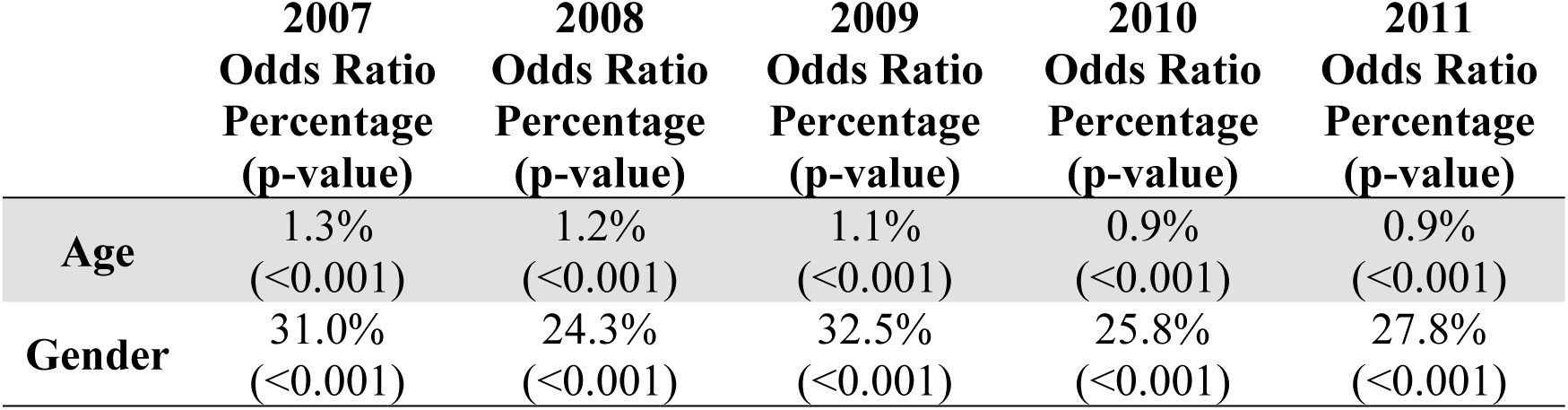
Benign Colorectal Neoplasms. The odds ratios (OR) percentage of having benign colorectal neoplasms only (reference= malignant or no neoplasms), using age and gender (reference=female) as independent variables. Furthermore, this data suggests that males have 24.3% - 32.5% higher odds of developing GI bleeding than females.

We then sought to determine whether the odds of lower GI bleed patients having benign or malignant colorectal neoplasms increases among patients who also have cardiovascular comorbidities. In 2010, patients with cardiovascular comorbidities had 10.5% (p= 0.04) higher odds of having benign colorectal neoplasms compared to those without cardiovascular comorbidities; however, results for the years 2007, 2008, 2009, and 2011, show no statistically significant difference in their odds of developing benign colorectal neoplasms between patients with and without cardiovascular comorbidities (Table 5). Conversely, for the years 2007-2011, the odds of malignant colorectal neoplasm diagnosis were 49.9%-65.3% higher for those without cardiovascular comorbidities, compared to those with cardiovascular comorbidities (Table 5). Lastly, among patients with lower GI bleeding, those without cardiovascular comorbidities had 49.7% to 75.1% higher odds of having malignant colorectal neoplasm diagnosis compared to those with cardiovascular comorbidities (Table 6).

**Table 5:** Colorectal Neoplasms and Cardiovascular Comorbidities. The odds ratio (OR) of having benign or malignant colorectal neoplasms (reference = no neoplasms) using cardiovascular comorbidities as an independent variable. This data suggests there is no statistically significant increase or decrease in the odds of developing a benign colorectal neoplasm, regardless of cardiovascular comorbidity status. Conversely, there is sufficient evidence to conclude the odds of being diagnosed with a malignant colorectal neoplasm after developing GI bleeding is 49.9-65.3% higher for patients without cardiovascular comorbidities. The highlighted values are those that are statistically significant.

|  | 2007<br>OR (p-value) | 2008<br>OR (p-value) | 2009<br>OR (p-value) | 2010<br>OR (p-value) | 2011<br>OR (p-value) |
| --- | --- | --- | --- | --- | --- |
| **Cardiovascular**<br>**Comorbidities and**<br>**Benign Neoplasms** | 0.905(0.06) | 0.985(0.80) | 1.045(0.38) | 1.105(0.04) | 1.012(0.82) |
| **Cardiovascular**<br>**Comorbidities and**<br>**Malignant**<br>**Neoplasms** | 0.605(<0.001) | 0.612(<0.001) | 0.632(<0.001) | 0.631(<0.001) | 0.667(<0.001) |

**Table 6:** Malignant Colorectal Neoplasms and Cardiovascular Comorbidities (when compared to Benign Neoplasms). The odds ratios (OR) of having malignant CRC (reference = benign neoplasm) using cardiovascular comorbidities as the independent variable. There is sufficient evidence to conclude the odds of patients admitted for GI bleeding having malignant CRC (as compared to benign colorectal neoplasms) are 49.7% (1/0.668=1.497) - 75.1% (1/0.571=1.751) higher for patients without cardiovascular comorbidities than for patients with cardiovascular comorbidities. The highlighted values are those that are statistically significant.

|  | 2007<br>OR (p-value) | 2008<br>OR (p-value) | 2009<br>OR (p-value) | 2010<br>OR (p-value) | 2011<br>OR (p-value) |
| --- | --- | --- | --- | --- | --- |
| **Cardiovascular**<br>**Comorbidities and**<br>**Malignant**<br>**Neoplasms**<br>**(Compared to**<br>**Benign)** | 0.668(<0.001) | 0.622(<0.001) | 0.605(<0.001) | 0.571(<0.001) | 0.659(<0.001) |

## Discussion

Male sex and increasing age are well established risk factors of colorectal neoplasms, which is confirmed in this study.^11–13^ According to our data and statistical analysis, patients without cardiovascular comorbidities have higher odds of being diagnosed with malignant colorectal cancer. Overall, patients admitted for lower GI bleed, regardless of cardiovascular comorbidities, shared similar rates of diagnosis with benign colorectal neoplasms (Table 5). There was a slight outlier in 2010, wherein patients with cardiovascular comorbidities presenting with lower GI bleeding had 10.5% higher odds (p=0.04) of a benign colorectal neoplasm diagnosis, as compared to those without cardiovascular comorbidities. While this is not sufficient data to support an overall increase in odds ratio, it does warrant further evaluation. Regarding malignant neoplasms, however, the data is clearer. Among patients with lower GI bleeding, patients without cardiovascular comorbidities have between 49.9% and 65.3% higher odds of being diagnosed with malignant colorectal neoplasms. We hypothesize that this may be partly due to earlier detection. Because patients with the selected cardiovascular comorbidities are highly likely to be taking anticoagulation or antiplatelet therapy, they are at higher risk of bleeding, and thus more likely to undergo colonoscopy and have benign neoplasms removed earlier, before they develop into malignant CRC. Notably, this cross-sectional study evaluates a possible association between CRC and cardiovascular comorbidities, using these diagnoses as a proxy for possible anticoagulation and antiplatelet therapies, but does not evaluate for causation. Therefore, future clinical trials are needed to determine how much earlier, within the CRC staging system, patients with cardiovascular comorbidities present with lower GI bleeding, and to investigate whether other factors contribute to their earlier presentation and more benign disease phenotypes.

The study population demonstrated notable demographic shifts over the study period from 2007 to 2011. The average age of patients remained stable, with no significant trend in mean or median values. However, there was a statistically significant increase in the proportion of patients aged 35-49 and 50–64 years, suggesting a growing burden among middle-aged groups, and highlighting the need for further evaluation of Early Onset Colorectal Cancer (EOCRC) risk among patients with cardiovascular comorbidities.

A known limitation of this study was that it included a slightly disproportionate number of patients identifying as White, as compared to patients identifying as Black or Hispanic.Minority populations, particularly Black and Hispanic individuals, may be disproportionately affected by comorbid conditions such as atrial fibrillation or cardiovascular disease, suggesting a possible increase in anticoagulant use and a subsequently higher risk of lower GI bleeding, thus further evaluation is needed to evaluate these populations. Paradoxically, this increase in bleeding events may present an opportunity for earlier CRC detection, as bleeding often prompts diagnostic evaluation such as colonoscopy. However, the effectiveness of this incidental screening pathway may be limited by disparities in follow-up care, healthcare literacy, and access to timely diagnostic services among minority patients.

Regarding income classifications, patients in the lowest income quartile comprised the largest socioeconomic group (average 30.03% from 2007–2011), highlighting the need for timely follow-up in underserved populations to prevent delays in malignancy detection. These demographic trends may reflect broader population-level changes in access to care and shifting healthcare utilization patterns, all of which are important considerations in interpreting disease burden and outcomes.

In addition to providing a basis for future clinical trials, the results from this study combined with other supporting studies can further influence recommendations regarding CRC screening intervals and frequency. Initiation of CRC screening at age 45 for asymptomatic, average risk adults may be adjusted according to the findings of this study, pendingfurther clinical investigation to establish causation.^22^ Although current colorectal cancer screening guidelines have become more stringent, there may be a need to reconsider screening strategies in patients with cardiovascular comorbidities who are on anticoagulants or antithrombotic therapy. These patients are at increased risk of lower GI bleeding, which may serve as an early warning sign for underlying colorectal pathology. Incorporating additional screening questions and adopting more proactive screening approaches—such as fecal immunochemical testing or early colonoscopy—may facilitate earlier detection of benign and malignant neoplasms. Tailoring screening protocols to account for bleeding risks in anticoagulated patients could improve outcomes without requiring universally more aggressive screening guidelines. Direct health policy and risk stratification may also be influenced by further evaluation of these results, with health systems and payers considering the incorporation of comorbidity status when triaging CRC screenings or prioritizing colonoscopy access.

This study is not without limitations. First, the NIS database does not collect information on outpatient medications. Therefore, there is no way to truly know in the absence of randomized clinical trials whether patients with cardiovascular comorbidities were on antiplatelet therapy and/or anticoagulation therapy. Additionally, many confounding variables such as family history of CRC, prior screening history, medication adherence, duration and dosage of anticoagulation and antiplatelet therapy, and lifestyle factors (smoking, physical activity, diet) are not available in the NIS database and may skew the findings. Furthermore, because NIS data is based on billing and administrative data for hospitalized patients, ICD-9 codes may not always accurately reflect clinical diagnoses or disease severity regarding CRC diagnoses and are specific to hospitalized patients. Therefore, the information may not be generalizable or applicable to the outpatient setting. Because this study utilized only hospitalization data, the exclusion of patients diagnosed in outpatient settings may further limit the generalizability to a broader CRC population. Furthermore, the regression analyses were only minimally adjusted, residual confounding cannot be excluded. Important demographic and socioeconomic factors such as race, insurance status, income level, and geographic region were not included in the regression models. Consequently, the observed associations should be interpreted as exploratory and hypothesis-generating. Due to the limitations of this cross-sectional study design, prospective studies and randomized controlled trials assessing CRC screening intervals in patients with or without cardiovascular comorbidities may provide more insight into early detection and outcomes related to CRC. Additionally, further research to stratify CRC diagnoses by stage at presentation can further validate the early detection hypothesis due to lower GI bleeding proposed in this paper.

The conclusions presented are based solely on the presence or absence of cardiovascular comorbidities. Medication-centered studies assessing the use of specific antiplatelet and anticoagulant regimes are also needed. Several studies have been done regarding the use of aspirin in CRC^23^, which dovetails well with this study. Specifically, in a large 2016 prospective cohort study of patients taking aspirin, the incidence of CRC was significantly lower compared to patients who were not taking aspirin. Although this study did not propose a mechanism, it did lead to several follow-up studies. More recent studies suggest aspirin use may be beneficial in CRC with specific tumor markers^24^, such as PI3K mutated tumors, while other studies suggest that all CRCs benefit from aspirin by mechanism of reduced inflammation^25^. While currently literature is still actively evaluating the efficacy and benefits of aspirin in CRC, this study encourages a broader look at CRC’s association with other anti-platelet and anticoagulant therapies and also contributes another proposed mechanism by which anticoagulation and anti-platelet therapy may contribute to a reduction in CRC rates. As with all cross-sectional studies, no causal inferences can be made regarding whether cardiovascular comorbidities directly influence potential cancerous neoplasm detection time or severity; however, this study supports the need for further prospective studies and randomized clinical trials.

## Conclusion

This manuscript analyzes NIS data from 2007 to 2011 and demonstrates that patients presenting with lower GI bleeding without cardiovascular comorbidities were significantly more likely to be diagnosed with malignant colorectal neoplasms compared to those with cardiovascular comorbidities. This information should initiate the discussion of whether patients without cardiovascular comorbidities should be screened for CRC differently, and *why* patients with cardiovascular comorbidities present for evaluation earlier or with more benign phenotypes. Clinical trials are warranted before clinical guidelines are changed; however, this manuscript can provide the basis for the conversation to optimize CRC screening and diagnosis.

## Data Availability

The data underlying the findings of this study are from the Healthcare Cost and Utilization Project (HCUP) National (Nationwide) Inpatient Sample (NIS), maintained by the Agency for Healthcare Research and Quality (AHRQ). The NIS is a third-party dataset available through the HCUP Central Distributor under a Data Use Agreement (DUA). The authors obtained and used the data in accordance with the HCUP DUA and all applicable publication requirements. The authors did not receive any special privileges in accessing the data beyond those available to any qualified purchaser. Researchers wishing to obtain the data may complete the required HCUP Data Use Agreement Training and purchase the NIS through the HCUP Central Distributor. Information regarding access, documentation, training, and purchasing is available through the HCUP website: https://hcup-us.ahrq.gov/db/nation/nis/nisdbdocumentation.jsp and https://hcup-us.ahrq.gov/tech_assist/centdist.jsp

https://www.hcup-us.ahrq.gov

## Ethical Approval

The involved institutions do not require ethical approval for NIS database studies.

